# Characterisation of Medication-Related Errors and Prioritisation of Patient-Safety Interventions in Hospital Care: A Retrospective Observational Study

**DOI:** 10.64898/2026.10.11.26365245

**Authors:** Chaitali Ravindra Kokate, Suman Muthumalli, Preethi Devarkonda, Niharika Jaiswal

**Affiliations:** Qualicare Healthcare Solutions.Nagpur, Maharashtra, India; Department of Pharmacology, IIAG Hospital Kozhikode, Kerelam, India; Aureus School of Pharmacy, Hyderabad, India; ACHS Hospital, Ratnagiri, MH, India

**Author notes:** **Ethical Committee Approval Information:** The protocol was reviewed and approved by the Ethical committee by Saraswati Mult speciality Hospital. **Funding:** This research received no specific grant from any funding agency in the public, commercial, or not-for-profit sectors. The study was conducted as an independent research project using routinely collected hospital medication-audit data.

**Keywords:** Medication-related incidents, Medication errors, Patient safety, Critical care, Inpatient care, Medication transcription errors, Medication omission, Medication administration, Root Cause Analysis, Pareto analysis, Poka-Yoke, Error-proofing, Quality improvement, Medication safety, PDSA

## Abstract

**Rationale:** Medication-related incidents are an important patient-safety concern, particularly in critical-care and inpatient settings where polypharmacy, clinical complexity, frequent therapeutic modifications, and transitions of care increase the risk of medication errors. Systematic analysis of reported incidents can help identify predominant error patterns, potential contributing factors, and priority areas for quality improvement. The present study was undertaken to characterize medication-related incidents and identify areas requiring targeted patient-safety interventions.

**Research Question:** What are the predominant patterns and distribution of medication-related incidents across critical-care and inpatient units, what patient-care, process, communication, documentation, and system-level factors potentially contribute to their occurrence, and how can these findings be systematically translated into prioritized, evidence-based quality-improvement interventions to strengthen medication safety and reduce preventable medication-related harm?

**Study Design and Methods:** A retrospective descriptive observational study was conducted to assess the pattern of medication-related errors identified through medication audits conducted in inpatient and critical-care settings. A total of 74 medication audit records were retrospectively reviewed. Each record was evaluated for the presence and type of medication-related error, including transcription, administration, documentation, and dispensing errors.

The identified medication errors were categorized based on the nature of the error documented in the audit records. As individual records could contain more than one type of medication-related error, the error categories were considered non-mutually exclusive. Consequently, the total number of error occurrences could exceed the number of records reviewed.

Descriptive statistical analysis was performed using frequency distributions and percentages to determine the relative occurrence of each medication-error category. Pareto analysis was subsequently performed to identify the predominant error categories and establish priorities for quality-improvement interventions. The Pareto analysis demonstrated that transcription and administration errors constituted the major contributors to the observed medication-related errors.

The identified error patterns were further evaluated using a process-based patient-safety approach to determine potential failure points within the medication-use process. Poka-Yoke (error-proofing) principles were applied to identify opportunities for preventing or detecting errors at the point of occurrence. Based on the findings, potential interventions included standardization of medication-ordering and transcription processes, mandatory verification of medication details, strengthening of independent verification for high-alert medications, and incorporation of technology-enabled safeguards such as electronic medication administration records and barcode-based medication administration.

The findings were used to identify priority areas for targeted quality-improvement interventions aimed at reducing medication-related errors and strengthening the safety and reliability of the medication-use process.

**Results:** A total of 91 medication-related error occurrences were identified across the 74 audit records reviewed. Transcription errors constituted the predominant error category (46; 50.5%), followed by administration-related errors (41; 45.1%). Documentation-related and dispensing-related errors accounted for 3 (3.3%) and 1 (1.1%) occurrences, respectively. Pareto analysis demonstrated that transcription and administration-related errors collectively accounted for 87 of the 91 identified occurrences (95.6%), indicating a substantial concentration of medication-related errors within these two categories. The cumulative frequency increased from 50.5% for transcription errors to 95.6% following inclusion of administration-related errors, while the remaining categories contributed only 4.4% of the total error occurrences. Accordingly, transcription and administration-related errors were identified as the principal priority areas for subsequent root cause analysis and quality-improvement interventions.

**Interpretation:** The findings demonstrate that transcription errors (50.5%) and administration-related errors (45.1%) were the predominant medication-related errors, collectively accounting for 87 of the 91 identified error occurrences (95.6%). Documentation-related errors constituted 3.3%, while dispensing-related errors accounted for 1.1% of the observed occurrences. Pareto analysis revealed a marked concentration of medication-related errors within the transcription and administration domains, indicating that these two categories represented the principal areas requiring focused quality-improvement interventions. Subsequent Root Cause Analysis (RCA) and Poka-Yoke-based error-proofing approaches were therefore directed toward identifying and mitigating process vulnerabilities within these high-frequency error domains.

## Background

Medication-related errors remain a persistent and potentially preventable patient-safety challenge across healthcare systems and constitute an important source of avoidable morbidity and mortality. Medication use is a complex, multistep process encompassing prescribing, transcription, dispensing, administration, monitoring, and documentation, with each stage presenting distinct opportunities for error. Failures within these processes may result in delays or omissions in appropriate treatment, inappropriate medication use, adverse drug events (ADEs), prolonged hospitalization, increased healthcare utilization, and preventable patient harm. The World Health Organization (WHO), through its Global Patient Safety Challenge: Medication Without Harm, has recognized medication-related harm as a major global patient-safety priority and has called for systematic action to reduce severe avoidable medication-related harm. WHO specifically identifies high-risk situations, polypharmacy, and transitions of care as priority areas requiring coordinated attention across patients, healthcare professionals, medicines, and medication-use systems.

Medication errors may occur at different stages of the medication-use pathway and frequently reflect interactions between human factors and weaknesses within healthcare systems. WHO recognizes that factors such as fatigue, inadequate working environments, staffing constraints, and weaknesses in medication systems can influence prescribing, transcribing, dispensing, administration, and monitoring practices. Importantly, medication-related errors should therefore not be conceptualized solely as individual failures but as events emerging from the interaction of healthcare professionals with complex clinical and organizational systems.

The WHO framework further emphasizes that medication safety requires interventions directed at both individual practices and the systems in which medicines are prescribed, prepared, administered, and monitored.

Within this medication-use pathway, transcription represents an important point of vulnerability, as medication information must be accurately transferred between prescribing and subsequent clinical documentation and administration processes. Errors introduced during this transition may propagate into downstream stages of medication management. Similarly, administration-related errors represent a critical stage because they directly influence whether the intended medication therapy is delivered accurately to the patient. The identification and characterization of these errors are therefore important not only for quantifying their frequency but also for understanding where within the medication-use pathway vulnerabilities are concentrated.

The Indian healthcare context further reinforces the importance of rational and safe medication use. The Indian Council of Medical Research (ICMR) provides evidence-based clinical and treatment guidance intended to support appropriate clinical decision-making, including guidance addressing antimicrobial use and antimicrobial stewardship. Such guidance reflects the broader principle that medication use should be evidence-based, clinically justified, and appropriately monitored. The Indian Medical Association (IMA) has also been involved in promoting medication safety through its collaboration with the Pharmacovigilance Programme of India (PvPI). The IMA-PvPI initiative emphasizes detection, assessment, understanding, and prevention of adverse effects and other drug-related problems, while promoting patient safety, rational use of medicines, and systematic reporting of adverse drug reactions. In addition, IMA’s published professional guidance emphasizes rational prescribing and appropriate use of medicines as an element of physicians’ responsibilities.

The importance of medication safety is also reflected in international recommendations addressing specific high-risk components of medication use. WHO identifies high-alert medications, provider and patient factors, and system or work-environment factors as important contributors to high-risk medication situations. Its medication-safety framework additionally emphasizes the importance of medication reconciliation and continuity of medication information during transitions of care, as well as systematic approaches to medication safety in patients exposed to polypharmacy. These recommendations highlight that medication safety is fundamentally a systems-level phenomenon requiring reliable processes across multiple stages of care rather than dependence on individual vigilance alone.

Despite increasing international and national emphasis on medication safety, the pattern and distribution of medication-related errors may vary substantially between healthcare institutions because of differences in patient populations, clinical complexity, medication-use systems, professional workflows, documentation practices, staffing patterns, and organizational processes. Consequently, institution-specific evaluation remains important for identifying the predominant forms of medication-related error occurring in routine clinical practice. Quantifying these patterns can provide empirical insight into the stages of medication management that are most vulnerable to error and can establish a foundation for subsequent investigation of the factors associated with their occurrence.

In the present study, 74 patient records were systematically reviewed, identifying 91 medication-related error occurrences. Transcription errors were the most frequently observed category, accounting for 46 occurrences (50.5%), followed by administration-related errors, accounting for 41 occurrences (45.1%). The predominance of these categories suggests that medication-information transfer and medication administration constitute important components of the medication-use pathway in which errors were frequently observed within the study setting. Importantly, the identification of these patterns provides descriptive evidence of the local medication-safety profile while also raising questions regarding the underlying clinical and system-level factors that may contribute to their occurrence.

A systematic characterization of medication-related errors is therefore relevant not only for estimating the burden of medication-safety events but also for understanding the distribution of errors across different stages of medication management. Such analysis may help identify recurring patterns, distinguish predominant from less frequently observed error types, and generate hypotheses regarding potential contributing factors. In this context, structured medication-error surveillance can serve as an important component of evidence generation for patient-safety research and can inform subsequent studies examining causal mechanisms and preventive strategies.

Accordingly, the present study aimed to characterize the pattern and distribution of medication-related errors identified through the systematic review of 74 patient records and to determine the predominant categories of medication-related errors observed within the hospital setting. By examining the frequency and distribution of medication-related error occurrences, the study seeks to provide institution-specific evidence regarding medication-safety vulnerabilities and contribute to the broader evidence base supporting safer medication-use practices in hospitalized patients.

## Methods

### Study Design and Setting

This study was conducted as a retrospective observational study of medication-related errors among hospitalized patients at Saraswati Mult speciality Hospital, a tertiary-care multispecialty hospital in location Ratnagiri, during August 2024 to November 2024 . The study utilized routinely documented clinical information to retrospectively identify and characterize medication-related errors occurring during the medication-use process.

The study was designed to quantify the burden of medication-related errors, describe their distribution according to error type, and identify the predominant error categories requiring targeted medication-safety interventions. The study was reported in accordance with applicable recommendations of the Strengthening the Reporting of Observational Studies in Epidemiology (STROBE) statement for observational studies.

The study protocol was reviewed and approved by Saraswati Mult speciality Hospital Ethics Committee Review Board. All data were deidentified before analysis, and confidentiality of patient information was maintained throughout the study.

### Study Population and Eligibility Criteria

The study population comprised hospitalized patients whose medical records were available for medication-safety assessment during the predefined study period. Records were eligible for inclusion when medication-related documentation was sufficiently complete to permit assessment of the medication-use process and identification of medication-related errors.

A total of 74 patient records met the study criteria and were included in the final analysis.

The patient record was the unit of inclusion, whereas the individual medication-related error occurrence was the unit of analysis for error-frequency estimation. This distinction was prespecified because a single patient record could contain more than one medication-related error. Accordingly, the number of observed medication-error occurrences was not restricted to the number of patients included in the study.

### Data Sources and Data Collection

Data were obtained through retrospective review of the electronic and/or paper-based medical records of eligible hospitalized patients using a predefined structured medication-safety data-collection instrument.

The data-collection instrument was designed to capture medication-related errors occurring during the medication-use pathway. Relevant medication documentation was reviewed, including medication prescriptions/orders, medication administration records, treatment sheets, nursing documentation, pharmacy documentation, and other medication-related clinical records, according to the availability of documentation within the study setting.

Each record was independently assessed against the predefined medication-safety criteria. A medication-related error was recorded when the documented medication-use process demonstrated a deviation from the prescribed, intended, established, or expected medication-use process, according to the operational definitions specified in the study protocol.

For each identified error, the following information was recorded: patient record identifier, medication-error category, stage of the medication-use process, and relevant supporting documentation. Multiple errors identified within the same patient record were recorded as separate error occurrences.

### Medication-Error Classification

Medication-related errors were classified according to a predefined classification framework established before analysis. The classification system included transcription errors, administration errors, omission errors, documentation errors, dispensing errors, and other predefined medication-related errors.

#### Transcription errors

were defined as discrepancies occurring during transfer or transcription of medication information between medication orders, treatment documentation, medication administration records, or other relevant medication-use documents.

#### Administration errors

were defined as deviations occurring during medication administration, including discrepancies involving the prescribed medication, dose, route, timing, or other predefined administration parameters.

#### Omission errors

were defined as failure to administer a prescribed medication when administration was indicated and no documented clinical reason for omission was identified.

#### Documentation errors

were defined as inaccuracies, omissions, or inconsistencies in required medication-related documentation that could affect the traceability or safety of the medication-use process.

#### Dispensing errors

were defined as discrepancies occurring during the medication-dispensing process, including provision of an incorrect medication, strength, quantity, or other dispensing parameter, where applicable.

The operational definitions were applied consistently during record review.

### Data Quality

To ensure the reliability of the extracted data, the completed audit records were reviewed for completeness, internal consistency, duplication, and appropriate categorization of medication-related errors before statistical analysis.

Where discrepancies were identified between the recorded error classification and the supporting clinical documentation, the source record was re-examined and the classification was resolved according to the predefined operational definitions. Duplicate entries referring to the same error occurrence were identified and removed where applicable.

The final dataset was subsequently locked for statistical analysis. The number of medication-related errors was determined from the finalized dataset rather than from preliminary audit counts.

### Study Outcomes

The primary outcome was the frequency and distribution of medication-related errors among the reviewed hospitalized patient records.

The primary outcome measures were:

1. The total number of medication-related error occurrences.
2. The frequency of each predefined medication-error category.
3. The percentage contribution of each error category to the total number of identified medication-related errors.
4. The cumulative percentage contribution of medication-error categories following frequency-based ranking.

The principal explanatory variable for the descriptive analysis was the type of medication-related error.

Where information was available and prespecified in the study protocol, additional variables including clinical department, medication class, medication-use stage, and other relevant patient or process characteristics were evaluated descriptively.

### Pareto-Based Prioritization of Medication Errors

To identify the medication-error categories contributing most substantially to the observed error burden, a Pareto analysis was performed.

Medication-error categories were ordered from highest to lowest frequency. The proportion contributed by each category was calculated relative to the total number of medication-related error occurrences, and cumulative percentages were subsequently derived.

The Pareto analysis was presented graphically using a Pareto chart, in which the frequency of individual error categories was represented on the primary axis and the cumulative percentage was represented on the secondary axis.

The purpose of the Pareto analysis was to identify high-contribution error categories for prioritization of medication-safety interventions. The commonly used 80% cumulative threshold was considered as a reference for prioritization; however, categories were not excluded from consideration solely on the basis of falling below this threshold. Clinical relevance, potential preventability, and feasibility of intervention were considered when translating Pareto findings into quality-improvement priorities.

#### Pareto Table.Table.01

| Rank | Medication-error category | Frequency (n) | Percentage (%) | Cumulative frequency | Cumulative (%) |
| --- | --- | --- | --- | --- | --- |
| 1 | Transcription errors | 46 | 50.5 | 46 | 50.5 |
| 2 | Administration errors | 41 | 45.1 | 87 | 95.6 |
| 3 | Other medication errors | 4 | 4.4 | 91 | 100.0 |
| **Total** | **All medication-related errors** | **91** | **100.0** | **91** | **100.0** |

### Mistake Proofing Framework

A Poka-Yoke-based error-proofing framework was used to identify potential process-level interventions for the prioritized error categories. Poka-Yoke was operationalized as the systematic identification of opportunities to prevent an error from occurring, make an error difficult to commit, or detect an error at an early stage before it could result in patient harm.

For each priority error category, the medication-use process was examined to determine whether the error could be addressed through standardization, simplification, forced verification, mandatory documentation fields, independent verification, standardized communication, visual differentiation, or other process controls.

The intervention framework therefore followed a sequential approach:

*Medication-error identification → frequency estimation → Pareto prioritization → process analysis → Poka-Yoke intervention design → corrective/preventive action*.

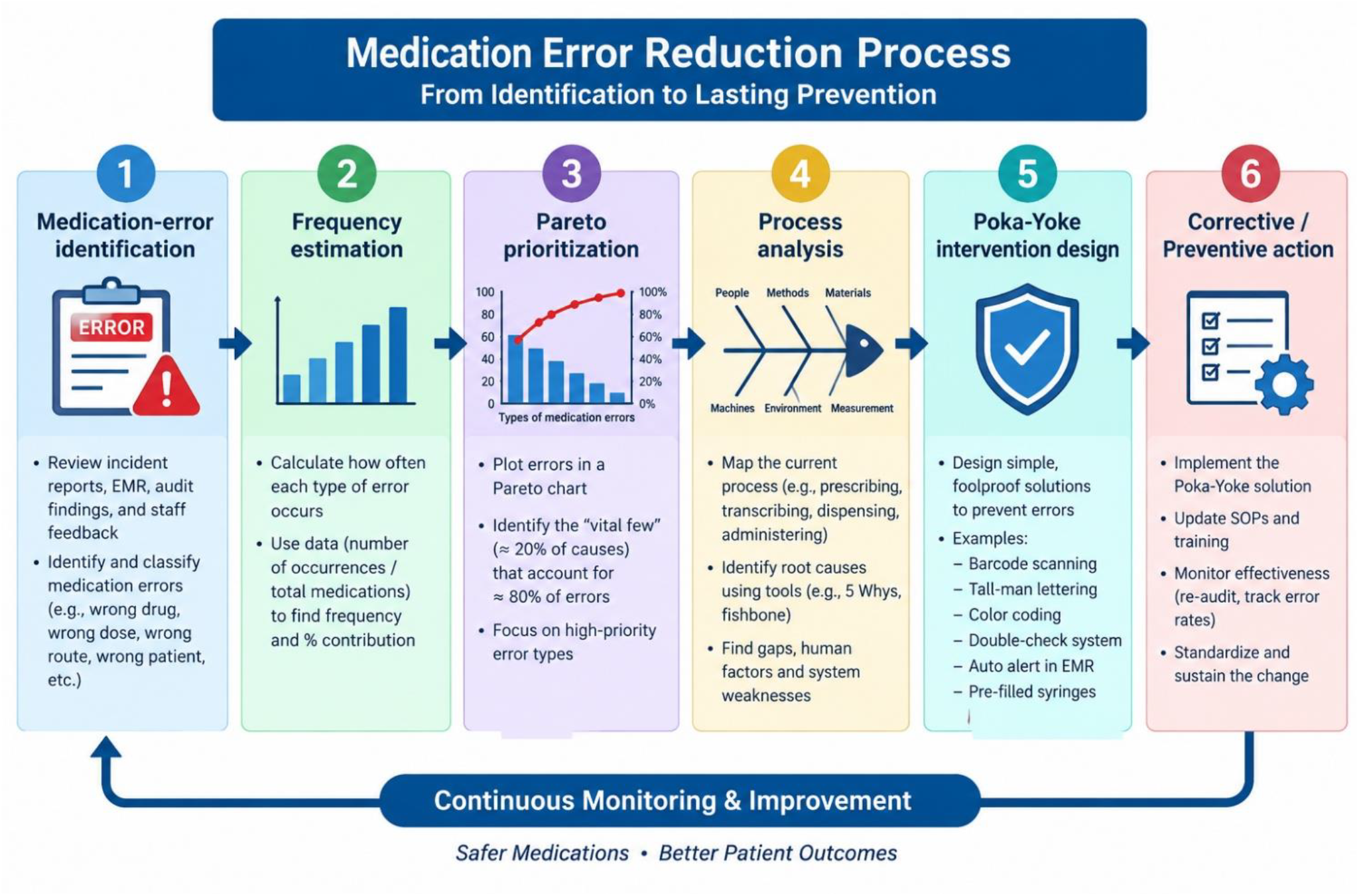

Poka-Yoke interventions were considered a quality-improvement component derived from the observational findings and were not treated as exposures in the retrospective observational analysis.

### Statistical Analysis

Statistical analysis was primarily descriptive. Categorical variables were summarized using frequencies and percentages. Continuous variables, where applicable, were summarized using appropriate measures of central tendency and dispersion.

A total of 74 patient records were reviewed. Across these records, 91 medication-related error occurrences were identified. The frequency of each medication-error category was calculated, and the percentage contribution of each category was determined using the total number of medication-related error occurrences as the denominator.

Among the identified errors, 46 (50.5%) were transcription-related and 41 (45.1%) were administration-related. The remaining 4 error occurrences were distributed among the other predefined medication-error categories.

Because the unit of analysis for error distribution was the individual error occurrence, rather than the patient record, the total number of error occurrences could exceed the number of patients included in the study.

Medication-error categories were subsequently ranked according to frequency and cumulative percentage for Pareto analysis. The resulting distribution was used to identify the categories contributing most substantially to the overall medication-error burden.

## Discussion

Medication-related errors represent a complex patient-safety problem arising from vulnerabilities at multiple points within the medication-use process. In the present retrospective medication-safety study, 74 patient records were reviewed and 91 medication-related error occurrences were identified. The occurrence of more errors than records reviewed indicates that medication-related failures were not necessarily isolated events and that multiple vulnerabilities could coexist within the same patient episode. The findings therefore support a systems-based approach to medication safety in which errors are examined not only in terms of their frequency but also in relation to the processes that permit them to occur.

### Predominant Medication-Error Patterns and Identification of Priority Problems

The analysis demonstrated that transcription errors were the predominant category, accounting for 46 of 91 error occurrences (50.5%), followed by administration-related errors, which accounted for 41 occurrences (45.1%). Together, these two categories constituted 87 of 91 identified medication-related errors (95.6%). This concentration indicates that the principal medication-safety vulnerabilities in the study setting were located at two critical interfaces: transfer of medication information and execution of medication administration at the point of care.

The predominance of transcription errors suggests that medication information may be vulnerable during transfer between prescribing, treatment documentation, medication administration records, and other clinical documentation systems. From a patient-safety perspective, transcription should not be considered merely an administrative activity. An inaccurate transcription can become an upstream failure capable of propagating through subsequent stages of the medication-use pathway. Consequently, a transcription discrepancy may remain undetected until medication administration, thereby creating the potential for downstream patient harm.

The second major problem identified was medication administration, which accounted for 45.1% of all error occurrences. Administration represents the final operational interface before medication reaches the patient and therefore constitutes an important safety barrier. Errors at this stage may arise from discrepancies in medication, dose, route, timing, patient identification, or other administration requirements. The substantial proportion of administration-related errors observed in this study indicates that strengthening bedside verification and standardization should constitute a major component of the medication-safety strategy.

### Pareto-Based Prioritization of the Medication-Safety Problem

The Pareto analysis provided an objective mechanism for translating the observed error distribution into improvement priorities. Transcription and administration errors accounted for 95.6% of all identified error occurrences, demonstrating that the majority of the observed medication-safety problem was concentrated within these two process categories. Therefore, rather than distributing corrective efforts uniformly across all medication-error categories, improvement resources should initially be directed toward the processes contributing to the greatest proportion of observed failures.

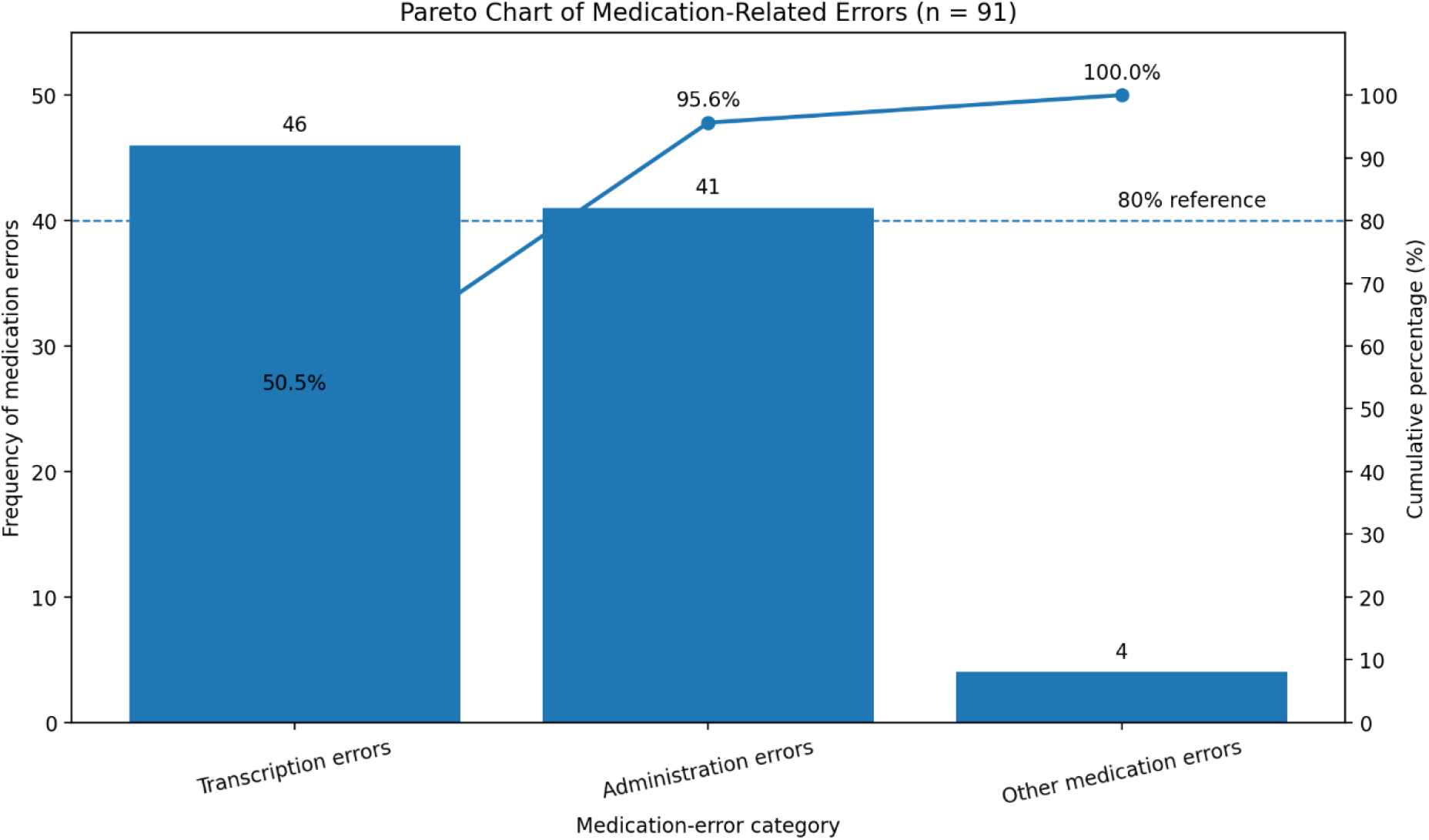

### Translation of Findings into Corrective and Preventive Action

The major contribution of the present analysis is the translation of error surveillance into a structured corrective and preventive framework. Identification of transcription and administration as the predominant error categories provides a specific basis for intervention rather than relying on generalized staff education alone.

For transcription errors, the identified process vulnerability can be addressed through standardized medication-transcription formats, reduction of unnecessary transcription points, structured medication reconciliation, mandatory verification of medication orders against the source prescription, and standardized documentation terminology. Where electronic systems are available, structured fields, alerts for discrepancies, and restrictions on incomplete medication entries may further reduce opportunities for transcription-related failure.

For administration-related errors, corrective strategies should focus on strengthening the final verification barrier before medication reaches the patient. Potential interventions include standardized medication-administration procedures, systematic verification of patient identity, medication, dose, route, timing, and relevant clinical parameters, barcode-assisted medication administration where available, and independent double-checking for selected high-risk medications. Standardized documentation immediately following administration may additionally improve traceability and facilitate early detection of discrepancies.

These interventions should be viewed as process controls rather than solely staff-dependent interventions. Education and competency assessment remain necessary components of medication safety; however, education alone may have limited effectiveness when the underlying process permits predictable errors. Accordingly, the findings support a transition from a predominantly person-centered approach toward a system-based medication-safety model.

### Application of Poka-Yoke to Medication-Safety Improvement

The findings provide a direct rationale for incorporating Poka-Yoke, or error-proofing, principles into medication-safety interventions. The objective of Poka-Yoke is not simply to remind healthcare professionals to avoid errors but to redesign the process so that an error is prevented, made difficult to commit, or detected before reaching the patient.

For transcription-related vulnerabilities, Poka-Yoke mechanisms may include mandatory fields for critical medication information, standardized electronic or paper transcription formats, automatic discrepancy alerts, restricted use of ambiguous abbreviations, and verification prompts requiring comparison between the original medication order and the transcribed record. Such interventions introduce additional system-level barriers against propagation of incorrect medication information.

For administration-related vulnerabilities, error-proofing may include barcode-based patient and medication verification, mandatory documentation of critical administration parameters, alerts for dose or route discrepancies, standardized high-alert medication checklists, and independent verification for medications associated with greater clinical risk. The underlying principle is to shift error detection as close as possible to the point at which the error is generated and, preferably, before medication administration occurs.

Thus, the proposed intervention pathway can be conceptualized as:

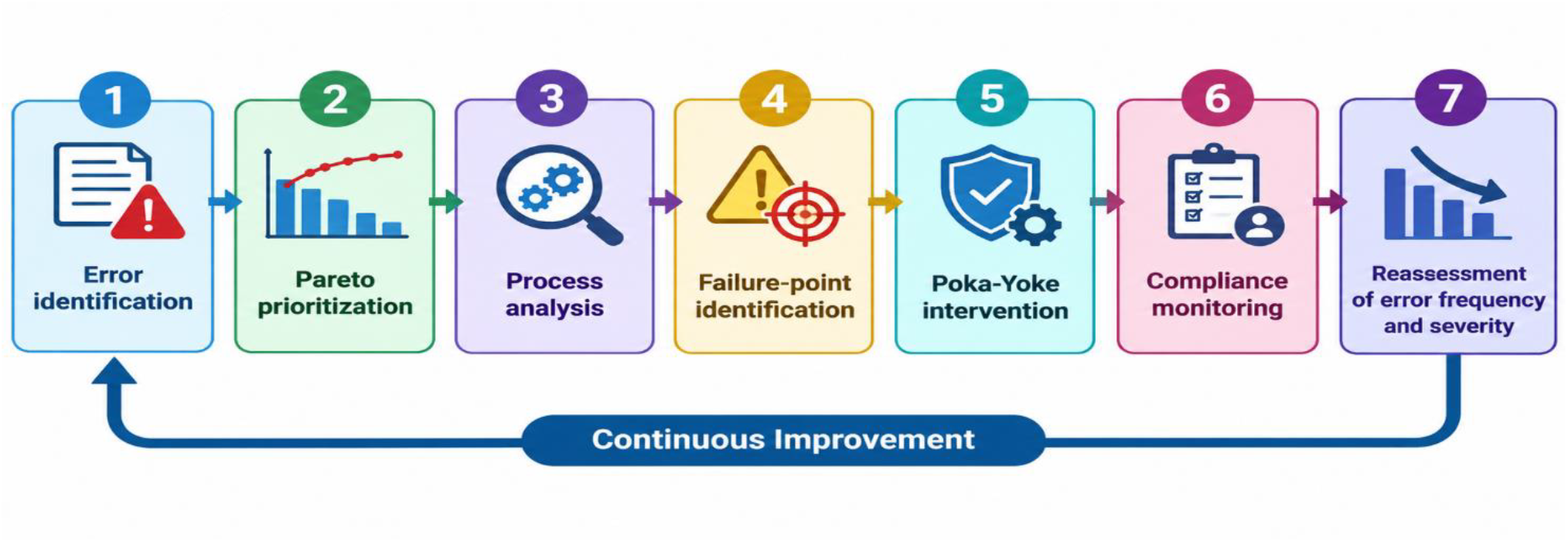

*Error identification → Pareto prioritization → Process analysis → Failure-point identification → Poka-Yoke intervention → Compliance monitoring → Reassessment of error frequency and severity*.

This framework provides a direct bridge between the observational findings and subsequent quality-improvement activity.

### From Detection to Prevention: Proposed Medication-Safety Model

The findings suggest that a sustainable medication-safety program should not terminate with identification and reporting of errors. A closed-loop improvement system should be established in which every major error category is linked to a defined preventive intervention, responsible process owner, monitoring indicator, and reassessment period.

For example, the high frequency of transcription errors should trigger review of the medication-information transfer pathway, identification of unnecessary or redundant transcription steps, implementation of standardized verification mechanisms, and subsequent measurement of transcription-error rates. Similarly, the high frequency of administration-related errors should lead to assessment of bedside medication-administration practices, identification of common failure points, implementation of standardized verification controls, and repeated monitoring of administration-error frequency.

Such an approach transforms medication-error surveillance from a passive reporting mechanism into an active patient-safety improvement system. Importantly, future assessment should determine whether the implemented interventions result in measurable reductions in error frequency, recurrence, severity, and preventability.

### Clinical and Quality-Improvement Implications

The findings have important implications for hospital medication-safety governance. The concentration of 95.6% of observed errors within transcription and administration processes suggests that targeted intervention may be more appropriate than broad, nonspecific medication-safety initiatives. Quality teams, pharmacy services, nursing leadership, and clinical departments can use these findings to jointly redesign vulnerable medication-use processes.

A multidisciplinary approach is particularly relevant because medication errors frequently occur across professional and departmental interfaces rather than within the isolated actions of a single healthcare professional. Accordingly, corrective actions should incorporate pharmacy, nursing, medical, information-technology, and quality-management perspectives where applicable.

The proposed interventions should also be accompanied by measurable indicators. These may include the medication-error rate per defined medication exposure, transcription-error rate, administration-error rate, compliance with medication-verification procedures, proportion of high-alert medications independently verified, and recurrence of previously identified error types. Monitoring these indicators over successive measurement periods would permit assessment of whether the interventions produce sustained improvement.

### Limitations

Several limitations should be considered when interpreting these findings. First, the retrospective observational design limits the ability to establish causal relationships between identified medication errors and patient outcomes. The presence of a medication-related error does not necessarily demonstrate that patient harm occurred, and the present study primarily assessed the frequency and distribution of errors rather than attributable clinical outcomes.

Second, retrospective analysis depends upon the quality and completeness of available documentation. Errors that were not documented or were inadequately recorded may not have been captured, potentially resulting in an underestimation of the actual medication-error burden. Third, the study was conducted within a single institutional setting, which may limit generalizability to hospitals with different patient populations, staffing models, medication-management systems, or levels of electronic integration.

Additionally, the Pareto analysis was based on error frequency and therefore does not necessarily identify the categories associated with the greatest severity or clinical harm. A relatively infrequent error may carry greater clinical consequences than a more frequently observed documentation-related error. Future analyses should therefore integrate frequency with severity, preventability, and potential patient harm.

Most importantly, the proposed Poka-Yoke and CAPA interventions should be considered planned or recommended improvement .The present study establishes the baseline problem and identifies priority intervention targets; it does not by itself demonstrate the effectiveness of the proposed interventions.

### Strengths

Despite these limitations, the study has several strengths. The analysis included 74 patient records and identified 91 medication-related error occurrences, permitting evaluation at the level of individual medication-error occurrences. The use of predefined error categories facilitated systematic classification, while Pareto analysis enabled prioritization of the dominant medication-safety problems.

A further strength is the integration of problem identification with a structured prevention framework. Rather than reporting error frequencies alone, the analysis identifies transcription and administration as the principal process vulnerabilities and provides a rationale for targeted CAPA and Poka-Yoke interventions. This creates a practical pathway from surveillance data to system redesign and subsequent reassessment.

## Conclusion and Future Direction

In this medication-safety study, transcription and administration-related errors constituted the predominant observed medication-error categories, together accounting for 95.6% of all identified error occurrences. The findings indicate that medication-safety improvement should initially focus on the processes responsible for medication-information transfer and bedside medication administration.

The study supports a structured improvement model in which Pareto analysis identifies priority problems, process analysis identifies contributory failure points, CAPA addresses the underlying process vulnerabilities, and Poka-Yoke mechanisms provide system-level error prevention or early detection. The effectiveness of this approach should subsequently be evaluated through prospective monitoring of medication-error frequency, severity, recurrence, preventability, and compliance with newly implemented safety controls.

Thus, the principal implication of the study is not merely that medication errors were identified, but that the observed error pattern provides an evidence-based foundation for targeted process redesign and measurable medication-safety improvement.

## Data Availability

All data produced in the present study are available upon reasonable request to the authors

https://www.who.int/publications/i/item/WHO-UHC-SDS-2019.11

## Acknowledgements

The authors would like to express their sincere gratitude to the management and healthcare professionals of Saraswati Multispeciality Hospital, Ratnagiri for their support and cooperation in facilitating this research. The authors acknowledge the contribution of the hospital’s medication-audit records, which provided the data required to analyse medication-related errors and identify opportunities for improving patient safety. The authors also appreciate the efforts of the healthcare professionals involved in medication management and documentation, whose routine clinical practices informed this study.

The authors are grateful to all individuals who contributed to the conceptualisation, data review, and preparation of this research manuscript.

